# Pulmonary Embolism Readmission Trend Over the Years: A National Readmission Database Study

**DOI:** 10.1101/2021.12.08.21267500

**Authors:** Mukunthan Murthi, Sujitha Velagapudi, Dae Yong Park, Hafeez Shaka

**Author notes:** Corresponding author: Mukunthan Murthi, Address: 1969, W. Ogden Ave, Chicago, IL-60612.

## Abstract

**Introduction:** Acute pulmonary embolism (PE) is known to be associated with significant short-term and long-term complications. However, with the evolution of PE management, the outcomes of PE-related complications and the need for readmission have not been well studied. The aim of this study is to see the trend in readmissions in PE patients from the years 2010 to 2018.

**Methods:** We utilized the National Readmission Database from 2010 to 2018 to identify hospitalized patients with a principal diagnosis of acute pulmonary embolism. Identified the total number of readmissions for acute PE from 2010 to 2018. A multivariate cox regression model was used to identify independent predictors of readmission.

**Results:** The number of patients with 30-day readmissions has gradually increased from 14,508 in 2010 to 19,703 in 2018. The proportion of females admitted was higher than males in all years. The 30-day all-cause readmission after principal admission for PE decreased from 11.2% to 9.7% from 2010 to 2014 but increased to 11.8% in 2018 (p<0.001). Risk-adjusted readmission specific for PE showed a decrease from 1.2 to 1% (p=0.023) from 2010 to 2018. When adjusted to age and gender, an increase in the proportion of readmissions with intracranial bleeding was seen among both the 30-day (0.7% in 2010 to 1.2% in 2018, aOR 1.06, p<0.001) and 90-day (0.7% in 2010 to 1.2% in 2018, aOR 1.06, p-trend 0.003) cohorts. Similarly, an increasing trend of readmissions for UGI bleed was seen among both 30-day (0.9% vs 4.3%, aOR=1.26, p-trend <0.001) and 90-day readmissions (0.7% vs 3.8%, aOR=1.27, p-trend <0.001). The units of blood transfusion required per readmission reduced in both cohorts during the study period.

**Conclusion:** Our study suggests that there is a statistically significant decrease in PE-specific readmission from 2010 to 2018, but an increase in all-cause readmissions. We also report an increase in non-major bleeding during readmissions, including ICH and UGI bleed. These findings warrant further studies to elucidate the mechanism for decreasing PE-specific readmission but possible causes for the increase in all-cause readmission in the hope of optimizing management and continuing improving outcomes.

## Introduction

As more sensitive laboratory testing and imaging studies emerge, more pulmonary embolism (PE) cases are being detected over the years (1). However, despite the increase in incidence, mortality has decreased from 7.1% in 1993 to 3.2% in 2012 (1). For most patients with PE who receive proper anticoagulation, outcomes are favorable, with the mean length of hospital stay also decreasing over time (2). Direct oral anticoagulants (DOAC) do not require titration of therapeutic levels facilitating early discharge of patients with the intent to continue anticoagulation at least for three months or longer depending upon PE etiology and risk factors (3). Because of these reasons, more patients with diagnosed pulmonary embolism are outside of the hospital.

With the increased detection and prevalence of pulmonary embolism, it is becoming more important to consider the post-hospital clinical course of patients with it. Especially with the Hospital Readmissions Reduction Program implemented by the Centers of Medicare and Medicaid Services, hospitals now need to reduce 30-day unplanned readmission rates after six conditions or specific procedures to avoid penalizations (4).

Owing to the relative commonality of venous thromboembolism (VTE), which includes both pulmonary embolism (P.E.) and deep vein thrombosis (DVT), a few studies in the past have examined the recurrence and causes of 30-day readmissions after VTE (5) (6). However, previous studies either looked at the recurrence of VTE as a whole or restricted the scope of readmission to within 30 days. Moreover, newer data has been issued over the years, providing more up-to-date statistics. As a result, this study aims to analyze national hospital readmission data in the United States (US) from 2010 to 2018 to discover the major causes of readmission after the diagnosis of pulmonary embolism until the wider timeline of 90 days, examine the characteristics of readmitted patients, and investigate how much these readmissions cause a burden on the healthcare system.

## MATERIALS AND METHODS

### Design and data source

This was a retrospective interrupted trends study involving adult hospitalizations for acute pulmonary embolism (PE) in the US. We extracted data from the Nationwide Readmissions Database (NRD) for 2010, 2012, 2014, 2016, and 2018. The NRD is the largest publicly available all-payer inpatient health care readmission database in the U.S., drawn from the Agency for Healthcare Research and Quality (AHRQ) Healthcare Cost and Utilization Project (HCUP) State Inpatient Databases (SID)(7). The NRD contains discharge data from geographically dispersed states within a calendar year. It contains both patient and hospital-level information. Hospitals are stratified according to ownership control, number of beds, teaching status, and metropolitan/non-metropolitan location. The NRD allows for weighted analysis to obtain 100% of the U.S. admissions within a given year. Databases before 2016 were coded using the International Classification of Diseases, Ninth Revision, Clinical Modification/Procedure Coding System (ICD-9-CM/PCS). Databases from 2016 were coded using the ICD-10-CM/PCS. In the NRD, diagnoses are divided into two separate categories: principal diagnosis and secondary diagnoses. A principal diagnosis was the ICD code attributed as the reason for hospitalization. Secondary diagnoses were any ICD code discharge diagnosis other than the principal diagnosis.

### Study population and variables

The study involved hospitalizations from each studied year of the NRD with APE as the reason for index admission using ICD codes (See supplementary table 1). We excluded hospitalizations with age less than 18, and elective hospitalizations. Using unique hospitalization identifiers, index hospitalizations were identified and one subsequent hospitalization within 30 or 90 days was tagged as readmission. Elective and traumatic admissions were excluded from readmissions. We excluded December admissions for the 30-day readmission analysis and October through December admissions for the 90-day readmission analysis as these hospitalizations would lack data for at least 30 days and 90 days respectively following the discharge to determine if there was readmission according to the study design. The NRD includes variables on patient demographics, including age, sex, median household income (income quartiles referred to patients as 1-low income, 2-middle income, 3-upper middle income, 4-high income), and primary payer. It also contains hospital-specific variables including bed size, teaching status, and location. We assessed the comorbidity burden using Sundararajan’s adaptation of the modified Deyo’s Charlson comorbidity index (CCI). This modification groups CCI into four groups in increasing risk for mortality (8). It has been adapted to population-based research to assess comorbidity burden. A score of >3 has about a 25% 10-year mortality, while a score of 2 or 1 has a 10% and 4% 10-year mortality, respectively. This cutoff point was chosen as a means of assessment of the increased risk of mortality (8). We obtained ICD codes for PE from literature review (9-11). The study analyzed validated diagnostic codes for intracranial hemorrhage (ICH), non-variceal upper gastrointestinal hemorrhage (NVUGIB), lower gastrointestinal hemorrhage (LGIB), and red blood transfusion from prior published literature (12-14).

### Outcome measures

We compared the top 10 reasons for both 30- and 90-day readmission following hospitalization for PE in 2010 and 2018. The study further highlights biodemographic trends in readmissions from 2010 to 2018. For both 30-day and 90-day readmissions, we calculated the yearly all-cause readmission rate, the PE-specific readmission rate, and the proportions of readmissions with any discharge diagnosis of ICH, NVUGIB, LGIB, and need for red blood cell transfusions. We also calculated the mean length of hospitalization (LOS), and inflation-adjusted mean total hospital cost (THC). The THC was obtained using the HCUP cost-to-charge Ratio files and adjusted for inflation using the Medical Expenditure Panel Survey index for hospital care, with 2018 as the reference point (15, 16).

### Statistical analysis

We used Stata® Version 16 software (StataCorp, Texas, USA) for the data analysis. All analyses were conducted using the weighted samples for national estimates in adjunct with HCUP regulations for using the NRD database. We employed multivariate logistic trend analysis to calculate risk-adjusted odds of trend in-hospital mortality, PE-specific readmission, ICH, NVUGIB, LGIB, and transfusion rates, mean LOS, and mean THC for readmissions using multivariate logistic trend analysis, adjusted for age and sex. All p values were 2 sided, with 0.05 set as the threshold for statistical significance.

#### Ethical considerations

The NRD database lacks patient identifiers. In keeping with other HCUP databases, the NRD database does not require Cook County Health Institutional Review Board approval for analysis.

### Data Availability Statement

The NRD is a large publicly available all-payer inpatient care database in the United States, containing data on more than 18 million hospital stays per year. Its large sample size provides sufficient data for analysis across hospital types and the study of readmissions for relatively uncommon disorders and procedures.

## RESULTS

### Patient demographics and hospital characteristics

During the study years, 721,710 patients were admitted with PE among which 80,014(11%) were non-electively readmitted within 30 days and 108,483(15%) were readmitted within 90 days of discharge (Figure 1). Patient demographics, comorbidities, and hospital characteristics are described in Table 1. The mean age of PE patients who got readmitted at 30- and 90-day were similar (63.2 vs 63.7 years). Females constituted the higher proportion of readmissions in both cohorts (54.2% vs 54.7%). More than half of the patients admitted were under Medicare payments (59.6% vs 60.8%). There was an increasing trend of admissions to metropolitan teaching hospitals at both 30-days (49.1% in 2010 to 73.4% in 2018) and 90-days (48.5% in 2010 to 72.8% in 2018). In both cohorts, more than 30% of the patients readmitted belonged to zip codes with a median annual household income of less than $28,999. The number of major medical comorbidities among readmitted patients gradually increased during the study period, with 36.1% patients in 2010 to 51.7% in 2018 having Charlson comorbidity index 3 or more. Similar trends were seen among the 90-day readmission cohort. (Table 2).

**Figure 1:**
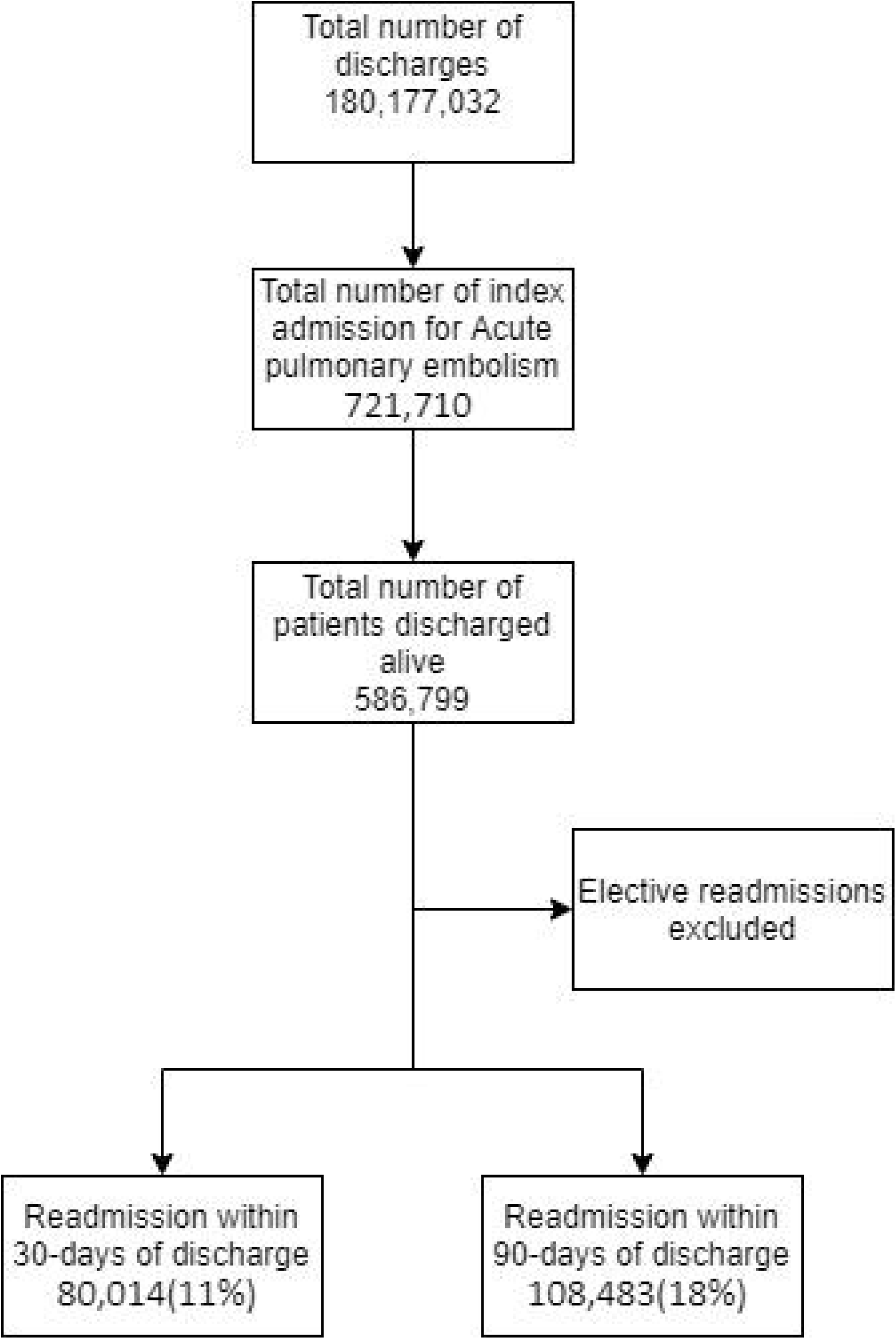
Flow chart showing patients selection for study

**Figure 2:**
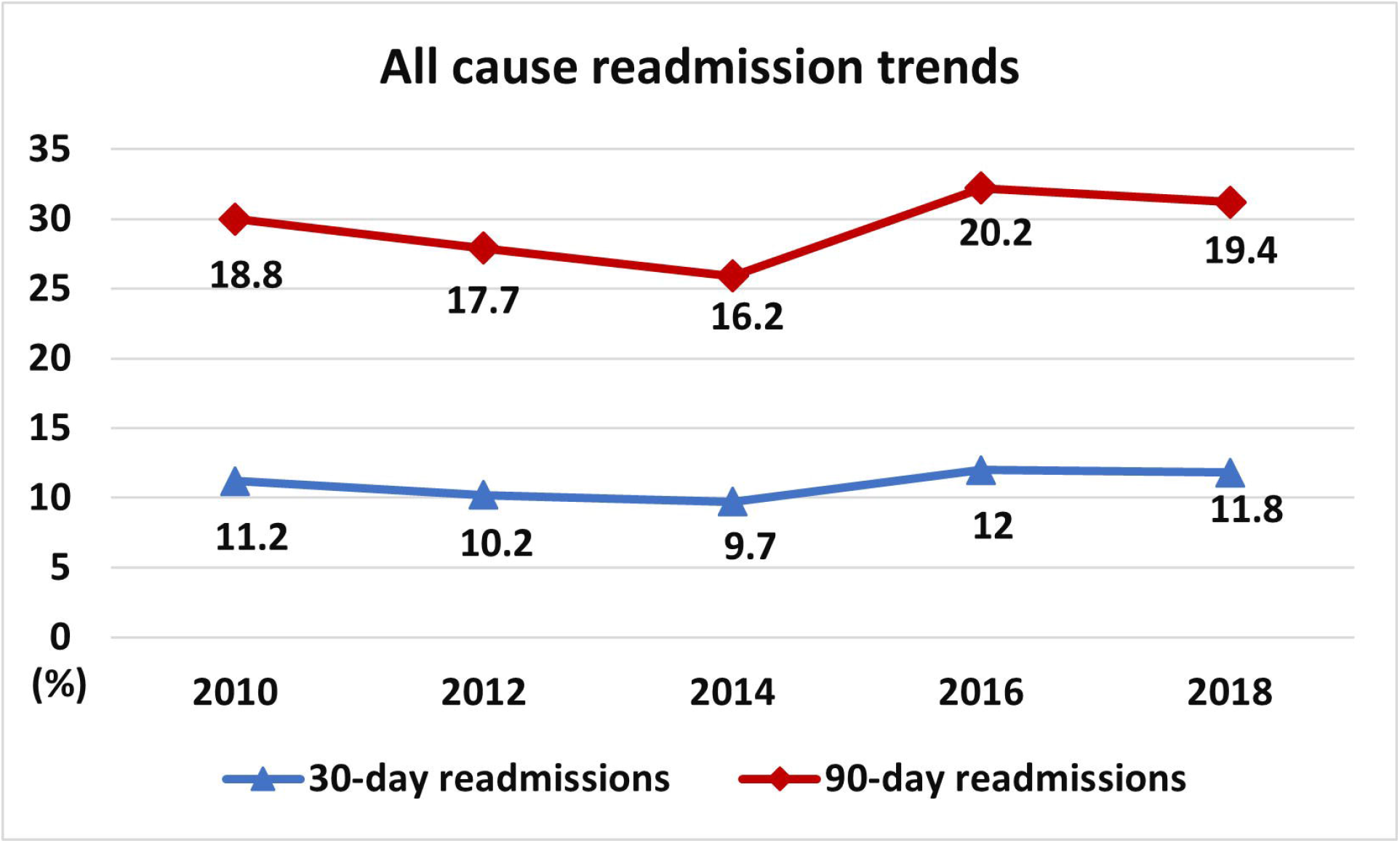
Graph showing all cause readmission rates after Pulmonary embolism from 2010 to 2018.

**Figure 3:**
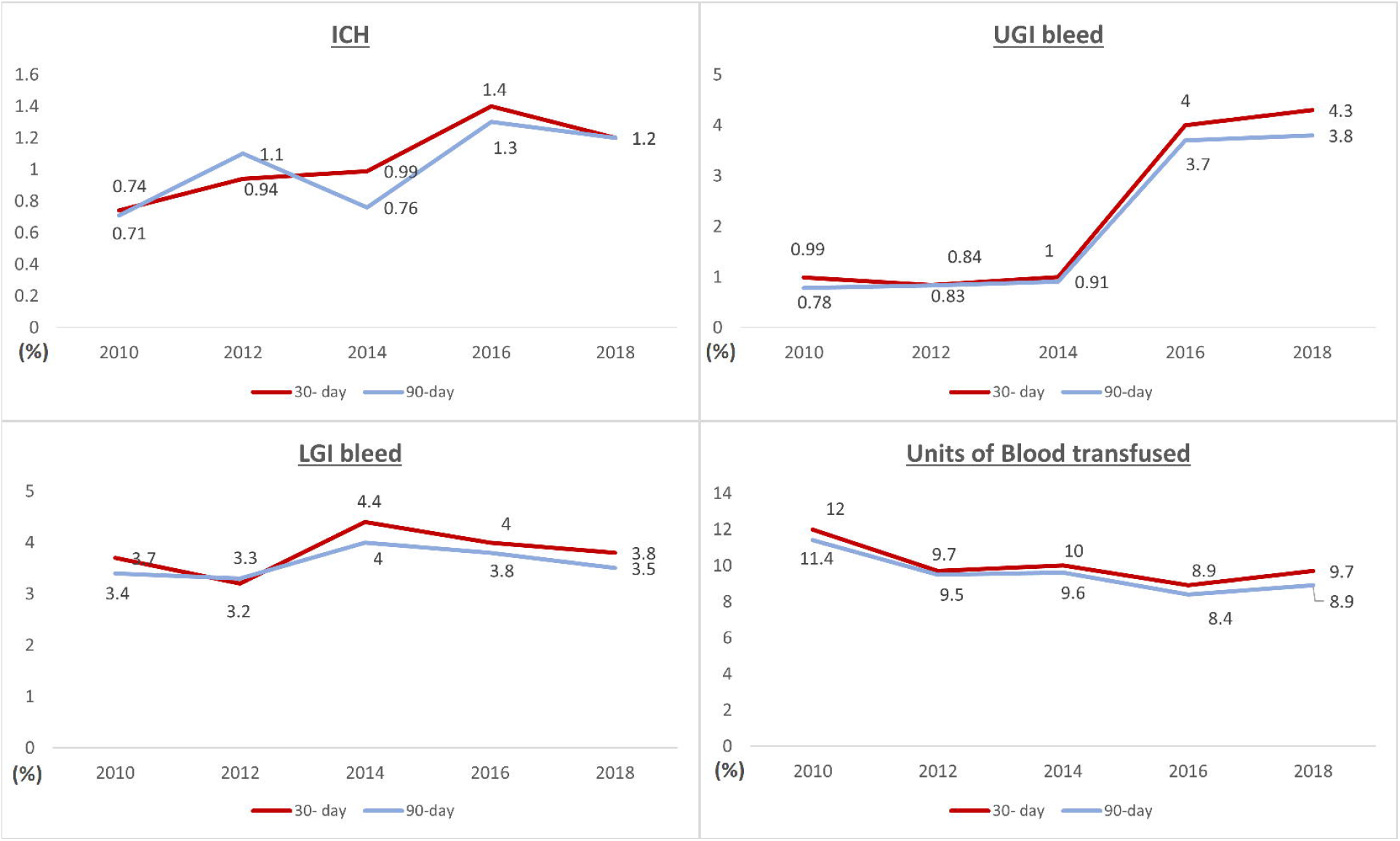
Rates of bleeding complications and blood transfusion requirements after PE readmission. ICH: Intracranial hemorrhage UGI: Upper Gastrointestinal LGI: Lower Gastrointestinal

**Table 1:** Trends in 30-days readmission after acute PE, patient demographics, hospital characteristics and in-hospital outcomes.

|  | **2010** | **2012** | **2014** | **2016** | **2018** | **p-value** |
| --- | --- | --- | --- | --- | --- | --- |
| **Readmitted (n)** | 14,508 | 13,882 | 13,283 | 19,024 | 19,317 | - |
| **Percentage readmitted** | 11.2 | 10.2 | 9.7 | 12 | 11.8 | <0.001 |
| **Mean age (years)(SE)** | 63.5±0.3 | 62.8±0.3 | 62.8±0.3 | 63.3±0.2 | 63.6±0.2 | - |
| **Female (%)** | 55.1 | 55.1 | 53.7 | 53.8 | 53.7 | 0.375 |
| **Charlson category 3 (%)** | 37.2 | 38.1 | 40.4 | 47.8 | 52.5 | <0.001 |
| **Insurance (%)** |  |  |  |  |  | 0.0009 |
| **Medicare** | 59.9 | 60.5 | 58.3 | 59 | 60.4 |  |
| **Medicaid** | 13.2 | 14.7 | 15.9 | 15.9 | 15.7 |  |
| **Private** | 22.9 | 20.6 | 21.8 | 22.1 | 20.4 |  |
| **Other** | 3.9 | 4.1 | 3.9 | 2.9 | 3.3 |  |
| **Zip-code wise median income (in \$) (%)** | | | | | | 0.021 |
| **1 - 45,999** | 32.4 | 33.4 | 31.1 | 31.7 | 31.2 |  |
| **46,000 - 58,999** | 25.1 | 25 | 27.9 | 25.5 | 28 |  |
| **59,000 - 78,999** | 22.4 | 22.2 | 22.1 | 24.7 | 24.1 |  |
| **79,000+** | 19.9 | 19.1 | 18.5 | 17.9 | 16.5 |  |
| **Hospital Bed-size (%)** |  |  |  |  |  | <0.001 |
| **Small** | 10.6 | 10.9 | 15.4 | 14.7 | 16 |  |
| **Medium** | 21.9 | 23.2 | 26 | 26.4 | 26 |  |
| **Large** | 67.4 | 65.7 | 58 | 58.8 | 57.9 |  |
| **Location/teaching status of hospital (%)** |  |  |  |  |  | <0.001 |
| **Rural** | 39.9 | 38 | 26.6 | 25.4 | 19.5 |  |
| **Urban non-teaching** | 49.1 | 50.8 | 64.7 | 66.3 | 73.4 |  |
| **Urban teaching** | 10.9 | 11.1 | 8.6 | 8.1 | 6.9 |  |
| **Readmission trends** |  |  |  |  |  | **p-value\*** |
| **PE (%)** | 1.2 | 1.2 | 1.1 | 1.1 | 1 | 0.004 |
| **ICH (%)** | 0.74 | 0.94 | 0.99 | 1.4 | 1.2 | 0.002 |
| **UGIB (%)** | 0.99 | 0.84 | 1 | 4 | 4.3 | <0.001 |
| **LGIB (%)** | 3.7 | 3.2 | 4.4 | 4 | 3.8 | 0.276 |
| **Blood transfusion (%)** | 12 | 9.7 | 10 | 8.9 | 9.7 | 0.002 |
| **Died (%)** | 6.5 | 6 | 6.5 | 6.4 | 6.9 | 0.357 |
| **Total cost (\$)** | 13,374 | 13,226 | 12,407 | 14,665 | 15,103 | <0.001 |
| **LOS (in days)** | 5.9 | 5.6 | 5.4 | 6 | 6.1 | 0.019 |
| **Total Length of stay (days)** | 86,222 | 78,379 | 72,078 | 114,502 | 117,866 |  |

\*Multivariate regression analysis

**Table 2:**
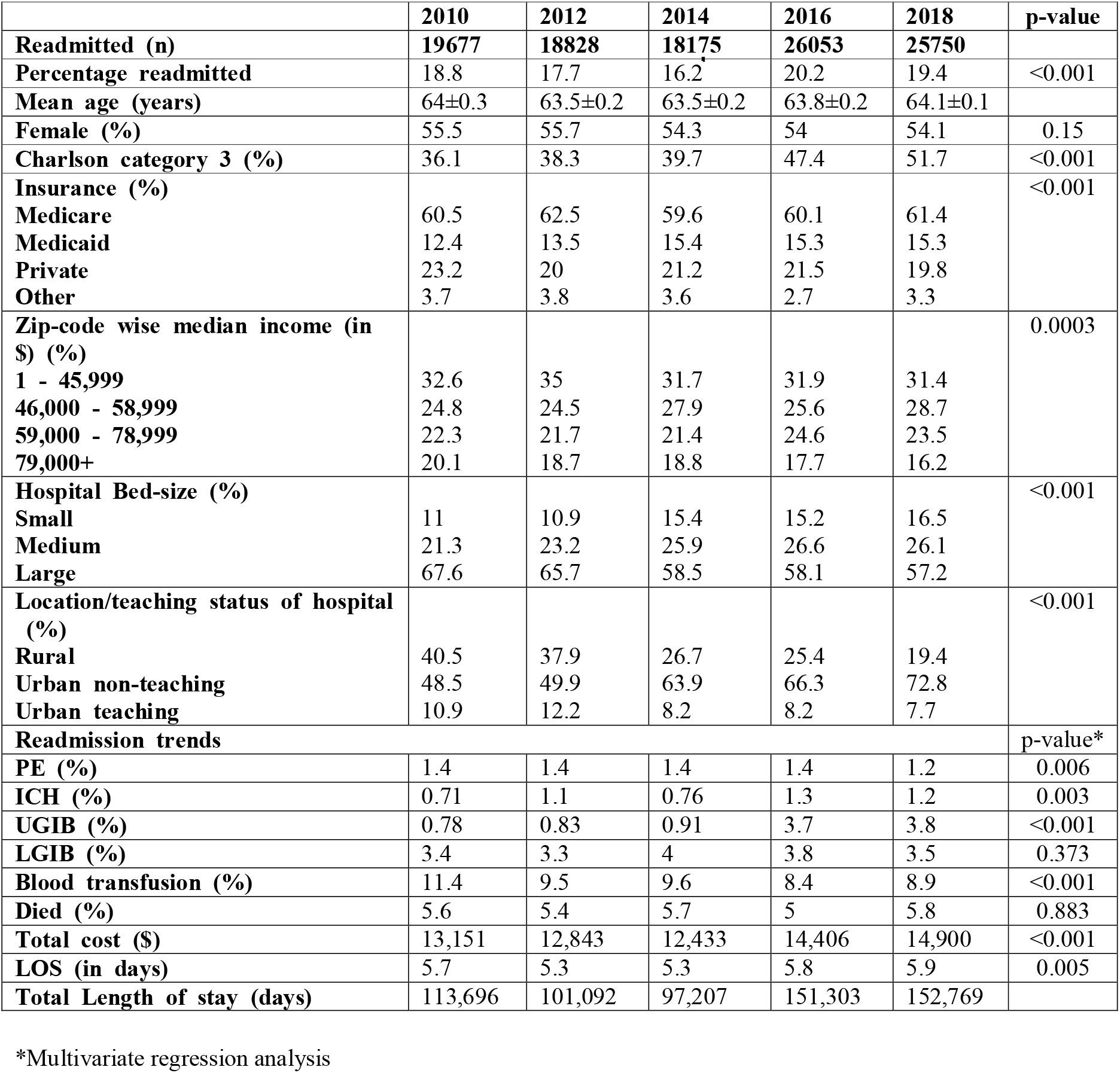
Trends in 90-days readmission after acute PE, patient demographics, hospital characteristics and in-hospital outcomes.

|  | **2010** | **2012** | **2014** | **2016** | **2018** | **p-value** |
| --- | --- | --- | --- | --- | --- | --- |
| **Readmitted (n)** | **19677** | **18828** | **18175** | **26053** | **25750** |  |
| **Percentage readmitted** | 18.8 | 17.7 | 16.2 | 20.2 | 19.4 | <0.001 |
| **Mean age (years)** | 64±0.3 | 63.5±0.2 | 63.5±0.2 | 63.8±0.2 | 64.1±0.1 |  |
| **Female (%)** | 55.5 | 55.7 | 54.3 | 54 | 54.1 | 0.15 |
| **Charlson category 3 (%)** | 36.1 | 38.3 | 39.7 | 47.4 | 51.7 | <0.001 |
| **Insurance (%)** |  |  |  |  |  | <0.001 |
| **Medicare** | 60.5 | 62.5 | 59.6 | 60.1 | 61.4 |  |
| **Medicaid** | 12.4 | 13.5 | 15.4 | 15.3 | 15.3 |  |
| **Private** | 23.2 | 20 | 21.2 | 21.5 | 19.8 |  |
| **Other** | 3.7 | 3.8 | 3.6 | 2.7 | 3.3 |  |
| **Zip-code wise median income (in \$) (%)** | | | | | | 0.0003 |
| **1 - 45,999** | 32.6 | 35 | 31.7 | 31.9 | 31.4 |  |
| **46,000 - 58,999** | 24.8 | 24.5 | 27.9 | 25.6 | 28.7 |  |
| **59,000 - 78,999** | 22.3 | 21.7 | 21.4 | 24.6 | 23.5 |  |
| **79,000+** | 20.1 | 18.7 | 18.8 | 17.7 | 16.2 |  |
| **Hospital Bed-size (%)** |  |  |  |  |  | <0.001 |
| **Small** | 11 | 10.9 | 15.4 | 15.2 | 16.5 |  |
| **Medium** | 21.3 | 23.2 | 25.9 | 26.6 | 26.1 |  |
| **Large** | 67.6 | 65.7 | 58.5 | 58.1 | 57.2 |  |
| **Location/teaching status of hospital (%)** |  |  |  |  |  | <0.001 |
| **Rural** | 40.5 | 37.9 | 26.7 | 25.4 | 19.4 |  |
| **Urban non-teaching** | 48.5 | 49.9 | 63.9 | 66.3 | 72.8 |  |
| **Urban teaching** | 10.9 | 12.2 | 8.2 | 8.2 | 7.7 |  |
| **Readmission trends** |  |  |  |  |  | p-value* |
| **PE (%)** | 1.4 | 1.4 | 1.4 | 1.4 | 1.2 | 0.006 |
| **ICH (%)** | 0.71 | 1.1 | 0.76 | 1.3 | 1.2 | 0.003 |
| **UGIB (%)** | 0.78 | 0.83 | 0.91 | 3.7 | 3.8 | <0.001 |
| **LGIB (%)** | 3.4 | 3.3 | 4 | 3.8 | 3.5 | 0.373 |
| **Blood transfusion (%)** | 11.4 | 9.5 | 9.6 | 8.4 | 8.9 | <0.001 |
| **Died (%)** | 5.6 | 5.4 | 5.7 | 5 | 5.8 | 0.883 |
| **Total cost (\$)** | 13,151 | 12,843 | 12,433 | 14,406 | 14,900 | <0.001 |
| **LOS (in days)** | 5.7 | 5.3 | 5.3 | 5.8 | 5.9 | 0.005 |
| **Total Length of stay (days)** | 113,696 | 101,092 | 97,207 | 151,303 | 152,769 |  |

\*Multivariate regression analysis

### All-cause and PE specific readmissions

The rate of adjusted all-cause 30-day readmission decreased initially from 2010 to 2014 (11.2% to 9.7%) followed by a subsequent increase in readmissions (11.8% in 2018) (p trend <0.001). We noted a similar trend in all-cause 90-day readmissions (Table 2). The rate of PE-specific readmissions showed a downward trend during the study period in both the 30-day (1.2% to 1%, aOR 0.98, p-trend 0.004) and 90-day cohort (1.4% to 1.2%, aOR 0.98, p-trend 0.006).

### Comparison between readmission in 2010 and 2018

Among readmitted patients, the most common cause of 30-day readmission in 2010 and 2018 was acute PE (11.9% vs 9.5%). The readmission percentage for sepsis doubled during the same interval (3.5% vs 7.8%). 1.8% of readmission in 2010 had acute DVT, making it the 6th most common cause of readmission. Interestingly less than 1% of readmissions in 2018 were due to acute DVT and it was not among the top 10 causes of readmission. Acute kidney failure (2.1%) was the fifth most common cause of readmission in 2018 which was not seen in 10 common causes of readmission in 2010. Of note, 2.6% were readmitted for intercostal pain in 2010 which was not seen in 2018. Among the 90-day readmission cohort, acute pulmonary embolism remained the most common cause of readmission (9.3%) in 2010 with sepsis being the most common cause in 2018(7.5%). The number of readmissions for PE reduced to 7.7% of all readmissions in 2018.

### Bleeding events

When adjusted to age and gender, an increase in the proportion of readmissions with intracranial bleeding was seen among both the 30-day (0.7% in 2010 to 1.2% in 2018, aOR 1.06, p<0.001) and 90-day (0.7% in 2010 to 1.2% in 2018, aOR 1.06, p-trend 0.003) cohorts. Similarly, an increasing trend of readmissions for UGI bleed was seen among both 30-day (0.9% vs 4.3%, aOR=1.26, p-trend <0.001) and 90-day readmissions (0.7% vs 3.8%, aOR=1.27, p-trend <0.001). We noted a downtrend in the number of units of blood transfusion required per readmission in both cohorts during the study period.

### Mortality, length of stay, and total cost

There were no significant changes in the mortality rate among 30-day (aOR 1.01, p-trend 0.357) and 90-day (aOR 0.99, p-trend 0.883) cohorts during the study period. The Length of stay for each admission was also similar over the years for both the cohorts. From 2010 to 2018, the mean total adjusted hospital charges per admission increased for both 30-day ($13,374 vs $15,103, p-trend <0.001) and 90-day ($13,151 vs $14,900, p-trend <0.001) readmissions.

## Discussion

Our study reports essential data such as 1) Steady downtrend in PE specific readmissions from the years 2010 to 2018 in both 30 days and 90day cohort 2) DVT is no longer among the ten most common causes of readmissions in 2018 in comparison to 2010 3) The percentage of readmission for sepsis doubled from 3.5 to 7.8% over the study period. 4) There is an increase in intracranial and upper G.I. bleed at both 30 days and 90days, with a downtrend in the required units of blood transfusions.

PE is one of the leading causes of cardiovascular death among hospitalized patients worldwide (17, 18). PE diagnosis and management have significantly improved over the last two decades(19). It is unclear if the advances in PE management, such as increasing direct-acting anticoagulant use, catheter-directed thrombolysis, and expanded use of inpatient and post-operative prophylaxis, have any mortality benefit. As per Karlyn et al., the age-adjusted mortality rate for PE has increased in the past decade (20). The increase in the burden of obesity has been hypothesized to contribute to poor cardiovascular outcomes, increasing PE-related mortality. However, Paul D stein et al. study demonstrated a decrease in the mortality in high-risk pulmonary embolism over the same time frame (21). This decrease in mortality was primarily attributed to improved shock and cardiac arrest management than advanced PE management strategies. Given this variability in in-hospital outcomes and mortality, our study mainly focuses on short-term and long-term rates of readmission in PE.

The 30-day readmission data mainly reflect the quality of acute care during the index hospitalization, safe care transitions, and immediate post-hospital period complications. The 90-day data is largely indicative of ambulatory management and care coordination. Our study demonstrated a downtrend in PE-specific readmissions at 30 days (1.2% to 1%) and 90 days (1.4 to 1.2%). One of the contributors could be the increase in DOAC usage. The proportion using DOACs has increased from 7.4% in 2011 to 66.8% in 2019, especially increasing among Medicare beneficiaries (22). DOACs are non-inferior to VKA (Vitamin-K antagonists) for VTE treatment as suggested by multiple Randomized control trials (RCTs) (23-25). As per the EINSTEIN-CHOICE, EINSTEIN-PE, and AMPLIFY trials, there was no difference in the rates of recurrent VTE with DOACs compared to VKA. This lack of difference in VTE recurrence was noted when the time in therapeutic range (TTR) for Warfarin is between 57 to 62%. However, it is unclear if this TTR is reflective of the general population as the subjects in the study have a closer follow-up. Hence, we hypothesize that DOACs are associated with a lower rate of VTE recurrence in the general population compared to VKA. This, along with improved care transitions could potentially explain the improved outcomes seen in our study.

DVT was the 6th most common cause of readmission in 2010. Interestingly it was not among the top ten causes of readmission in 2018. While the increase in DOAC usage, as mentioned above, could be contributing to the trend, the policy-directed approach to prevent VTE is the more likely contributing factor. To prevent VTE and care of patients with VTE, the National Quality Forum endorsed six VTE core measures in 2006 (26). VTE thromboprophylaxis among hospitalized patients was one of the critical components. In addition to this, postprocedural VTE listing under the hospital-acquired condition reduction program, tracking of VTE prophylaxis prescription by National Quality Forum, and requirement of centers of Medicare and Medicaid to report surgical care improvement project measures related to VTE are proposed to have led to an increased trend in thromboprophylaxis utilization (27, 28). The decrease in readmissions secondary to DVT could be a product of the above efforts. Additionally, the proportion of readmissions secondary to sepsis has doubled during the study period (3.5 to 7.8%). This uptrend is consistent with the increased incidence of sepsis due to increased clinical awareness, more screening, and decreased diagnostic thresholds, as demonstrated by numerous studies(29, 30).

The adjusted all-cause 30-day readmission decreased initially from 2010 to 2014 (11.2% to 9.7%), followed by a subsequent increase in readmissions (11.8% in 2018) (p trend <0.001). The initial decreasing trend is consistent with the 30-day readmission results in Behnood et al. study(31). This downtrend was thought to be secondary to improved PE management strategies. However, the reason for the subsequent uptick in readmissions remains unclear. One possible explanation could be the increasing comorbidity burden among PE patients, placing them at increased risk of readmission. Another potential cause for this upward trend could be the change in the coding system from ICD-9 to ICD-10 in the latter half of 2015.

Our study noted an increase in intracranial and UGI bleeding in 30 and 90-days, with a parallel decrease in the units of blood transfusions required in both cohorts. The above results could indicate the increase in clinically relevant non-major bleeding not requiring blood product transfusion. Apixaban (25%) is the most widely used DOAC, followed by rivaroxaban (21%)(32). The EINSTEIN VTE study showed an increased risk of clinically relevant non-major bleeding (mainly mucosal bleed) with rivaroxaban and no increase in major bleeding(33). Thus, widespread usage of DOAC could explain the trend of increasing non-major bleeding.

## Conclusion

Our study results are based on the one of the largest population databases. Our results show a steady downtrend in PE specific readmissions possibly due to the multitude of advancements in PE diagnosis and management over the decades, including increasing DOAC usage. Our data also shows reduction in the rates of major bleeding, evidenced by reduced blood transfusions in readmitted patients over the study period. Given the limited availability of laboratory and pharmacological data in NIS, further prospective studies are required to fully understand and confirm this trend.

## Data Availability

All data produced in the present work are contained in the manuscript

https://www.hcup-us.ahrq.gov/nisoverview.jsp

