## Supplemental Table 1 for "Pulmonary Embolism Readmission Trend Over the Years: A National Readmission Database Study"

| **Diagnosis** | **ICD-9 code** | **ICD-10 code** |
| --- | --- | --- |
| **Acute Pulmonary Embolism** | 41519 | I2692, I2693, I2694, I2694, I2699, I2602, I2609 |
| **Intracranial Hemorrhage** | 430, 431, 4320, 4321, 4329 | I60, I61, I62 |
| **Upper Gastrointestinal Bleeding** | 53100,53101,53120,53121,53220,53201,  53221,53300,53320,53301,53400,53401,  53420,53421,53501,53561, 5780,53784,  53021, 5307,53082 | K920, K921, K2210, K2212, K252, K260, K262, K270, K272, K280, K282, K2901, K2921, K2981, K31819, K3182 |
| **Lower gastrointestinal Bleeding** | 5781, 5789,5693 | K5521, K625, K922 |
| **Blood transfusion** | 9904 | 30230N0, 30230N1, 30230P0, 30230P1, 30233N1, 30233P0, 30233P1, 30240N0, 30240N1, 300240P1, 30243N0, 30243N1, 30243P0, 30243P1 |

**Supplementary table 1: List of ICD codes**
